# Association Between Somatic *PIK3CA* Mutations and Ancestry in South Asia: Prognostic Insights From a Sri Lankan Breast Cancer Study

**DOI:** 10.1101/2024.10.22.24315930

**Authors:** Tharini Ruwinya Cabraal, Iranthi Kumarasinghe, Ranga Perera, Jayantha Balawardane, Sameera Viswakula, Nandika Perera, Gayani Harendra Galhena

## Abstract

**Background:** The *PIK3CA* oncogene is one of the most mutated oncogenes in breast cancer, with ancestry-linked variations reported globally. The study aimed to discern the prevalence and prognostic role of *PIK3CA* mutations in Sri Lankan breast cancer patients for the first time, focusing on the correlation between somatic mutations and patient ancestry in an exclusively South Asian cohort of breast cancers.

**Materials & Methods:** A qPCR-based genetic analysis was performed on DNA from formalin-fixed, paraffin-embedded (FFPE) tissue samples of 63 clinically diagnosed, female, Sri Lankan breast cancer patients using the QClamp® *PIK3CA* Mutation Detection Test for the hotspot mutations of *PIK3CA* (i.e., H1047R, E545K, E542K) followed by a statistical analysis. Patient samples and clinical data were fully anonymized, with no identifying information available to authors at any point of the study.

**Results:** Somatic missense *PIK3CA* mutations H1047R and E542K, were detected in 17.46% of the cohort. E545K mutation was not detected. The observed mutations were associated with an increased risk of lymph node (LN) metastasis (p=0.036, OR 9.60) and reduced recurrence-free survival (RFS) (p<0.001, HR 26.19). In addition, LN metastasis (p=0.026, HR 123.94) and a high Ki67 index (p=0.029, HR 79.69) were individually associated with reduced RFS. All three factors above-presence of a *PIK3CA* mutation, LN metastasis and a high Ki67 index-in combination, were also associated with reduced RFS (p<0.001). Further analysis revealed a significant association between patient ancestry and *PIK3CA* mutation status (p=0.028).

**Conclusion:** Despite being a pilot study, the findings suggest that *PIK3CA* may serve as a potential prognostic biomarker in Sri Lankan breast cancer patients, with ancestry-linked variations potentially influencing metastatic outcomes. These results bring out the importance of integrating PI3K inhibitors into the therapeutic management of breast cancers in Sri Lanka after validating these findings in a functional study using a larger cohort.

## Introduction

Breast cancer is the most commonly diagnosed cancer in women worldwide [1]. Asian women, particularly those in developing countries, often have a poorer prognosis for breast cancer, with higher cancer grades and stages as well as an earlier onset of the disease [2].

Breast cancer has distinct molecular subtypes that are categorized based on the immunohistochemical (IHC) expression of steroid hormone receptors (SHR), including estrogen receptor (ER), progesterone receptor (PR), and human epidermal growth factor receptor 2 (HER2/*ERBB2* or HER2/*neu*). These subtypes are Luminal A, Luminal B, HER2-positive (HER2+), and triple-negative breast cancer (TNBC) [1].

More than 85% of breast cancers are sporadic, primarily caused by somatic mutations [3]. These mutations in oncogenes (OGs) and tumor suppressor genes (TSGs) often initiate tumorigenesis [4]. Breast cancer tumorigenesis has been linked to mutations in several cell signaling pathways crucial for normal development such as the PI3K/AKT/mTOR pathway, commonly referred to as the PI3K pathway. Genetic hyperactivation of this pathway is one of the most common driver mechanisms in breast cancer, often through alterations to its components like p110α (encoded by the phosphatidylinositol-4,5-bisphosphate 3-kinase catalytic subunit alpha gene/*PIK3CA; NCBI Gene ID 5290*), p110β, and p85α [1,5,6].

Approximately 30% of the mechanisms that over-activate PI3K signaling are due to somatic mutations in the *PIK3CA* oncogene [5], making it the most commonly mutated oncogene in breast cancer [6–8]. While *PIK3CA* mutations can occur across the p110α subunit, about 80% are caused by the three hotspot mutations clustered in two domains of p110α: E545K and E542K in the helical domain encoded by exon 9, and H1047R in the kinase domain encoded by exon 20. Among these, H1047R is the most frequently detected *PIK3CA* mutation, while E542K is the least prevalent [9].

Studies have shown that between 10 to 45% of all human breast cancers harbor *PIK3CA* driver mutations, with the exact percentage varying based on the studied population and ancestry [5–10]. Activating mutations of *PIK3CA* are reported in nearly 42% of SHR+/HER2-, 31% of HER2+ and 16% of TNBC tumors [5].

Importantly, the presence of *PIK3CA* mutations has been associated with poorer prognostic outcomes, especially in ER+ cases [11]. Numerous studies illustrate that *PIK3CA* mutations are linked with resistance to various therapies, including endocrine therapy, chemotherapy, radiotherapy, anti-HER2 therapy and immunotherapy. This resistance negatively affects all available treatment modalities for all breast cancer subtypes, with more pronounced effects in advanced and metastatic cases [5,7–10,12]. These mutations also promote cell division and inhibit apoptosis in TNBC cells, leading to resistance to their primary treatment option: chemotherapy [12]. In addition to their role in treatment resistance, accumulating evidence suggests a strong association between *PIK3CA* mutations and tumor recurrence, poor relapse/recurrence-free survival (RFS), and/or overall survival (OS), particularly in postoperative Asian patients with metastatic SHR+/HER2-breast cancers [9,11,13–15]. Despite extensive research, the overall relationship between *PIK3CA* mutations and the clinicopathological profile of breast cancer patients remains controversial, with conflicting findings across various patient populations, such as observed associations of *PIK3CA* mutations with longer RFS and/or OS [16,17].

Recent advances in genomic studies have provided significant insights into the relationship between genetic ancestry and somatic mutations across cancer types via changes in the germline [18–22]. In particular, the *PIK3CA* oncogene is reported as one of the top two genes observed in breast cancers, whose somatic mutations have shown significant ancestry-linked differences in several global studies predominantly involving patients from European and African ancestries [19–21]. However, Asian populations, particularly those of South Asian descent, remain highly underrepresented in these studies, limiting a comprehensive understanding of ancestry-driven somatic mutations in the region [22].

Sri Lanka is a unique island nation in the Indian Ocean with a rich tapestry of ethnic diversity. Over 70% of the population is composed of Sinhalese, descendants of Indo-Aryan ancestry, found exclusively in Sri Lanka [23]. Other major ethnic groups include Tamils and Moors, who trace their roots to Dravidian ancestry, and Burghers, descendants of European colonists in Sri Lanka, who also share Indo-Aryan ancestry [23,24]. This distinct combination of Indo-Aryan and Dravidian ancestries, together with its centralized geographical location, particularly during the maritime silk route, makes Sri Lanka a melting pot of genetic diversity.

Breast cancer is the most frequently diagnosed cancer among Sri Lankan women [25]. As a developing, lower-middle income island nation in the Indian Ocean, Sri Lanka has limited research on breast cancer trends and genetic markers, despite a significant rise in incidence and mortality rates over time [26,25]. To the best of our knowledge, no studies have been conducted on the prevalence and prognostic outcomes of *PIK3CA* mutations and their correlations with ancestry in breast cancer patients of Sri Lanka. Given the mutational spectrum of *PIK3CA* and its impact on the clinicopathological profile and prognosis of breast cancer, which varies significantly depending on the population and their ancestry, it is important to elucidate the current status of Sri Lankan breast cancer patients in this context – an area that remains largely unexplored.

This study was therefore designed to achieve the overarching aim of cataloguing hotspot mutations in the *PIK3CA* oncogene within a cross-section of Sri Lankan breast cancer patients that includes all different ancestral groups. Using DNA extracted from tumor tissues obtained at the point of diagnosis, we aimed to correlate the detected mutations with specific clinicopathological characteristics and ultimately assess the prognostic potential of *PIK3CA* mutations using RFS as a key metric. Given that the Sinhalese population is found nowhere else in the world (except for few migrant individuals), results from this study offer a rare opportunity to explore how ancestral genetic makeup in an island nation intersects with somatic mutations, such as those in the *PIK3CA* oncogene. This study managed to successfully catalog *PIK3CA* mutations in our cohort of Sri Lankan breast cancer patients and deduced their impacts on disease prognosis while addressing the critical gap on the implications of ancestry on somatic mutations in a purely South Asian cohort of breast cancer patients, for the first time.

## Materials and methods

### Patient samples

Ethical approval for the analysis of an existing set of formalin-fixed paraffin-embedded (FFPE) breast tumor biopsy samples and fully anonymized medical records of the relevant patients was obtained from the Ethics Review Committee of the Institute of Biology, Sri Lanka (ERC IOBSL 303 07 2023, S1 Fig). This retrospective study was conducted in compliance with the REMARK (Reporting Recommendations for Tumor Marker Prognostic Studies) and ICMJE (International Committee of Medical Journal Editors) guidelines.

The fully anonymized database of breast cancer patient records from the Diagnostics Laboratory at the University Hospital of General Sir John Kotelawala Defense University was accessed on the 27^th^ of August 2023. All patient data were fully anonymized, ensuring no patient identifiers were available to the authors at any stage of the research. Using this database, medical records of breast cancer patients treated at the University Hospital of General Sir John Kotelawala Defense University from 2019 to 2023, who met the inclusion criteria (Fig 1) were selected for the study. The corresponding FFPE blocks were retrieved from the hospital’s diagnostics laboratory and stored at room temperature until further analysis. Clinicopathological data and follow-up information of these patients ranging from 1 to 5 years post-diagnosis were collected from the hospital database.

**Fig 1.**
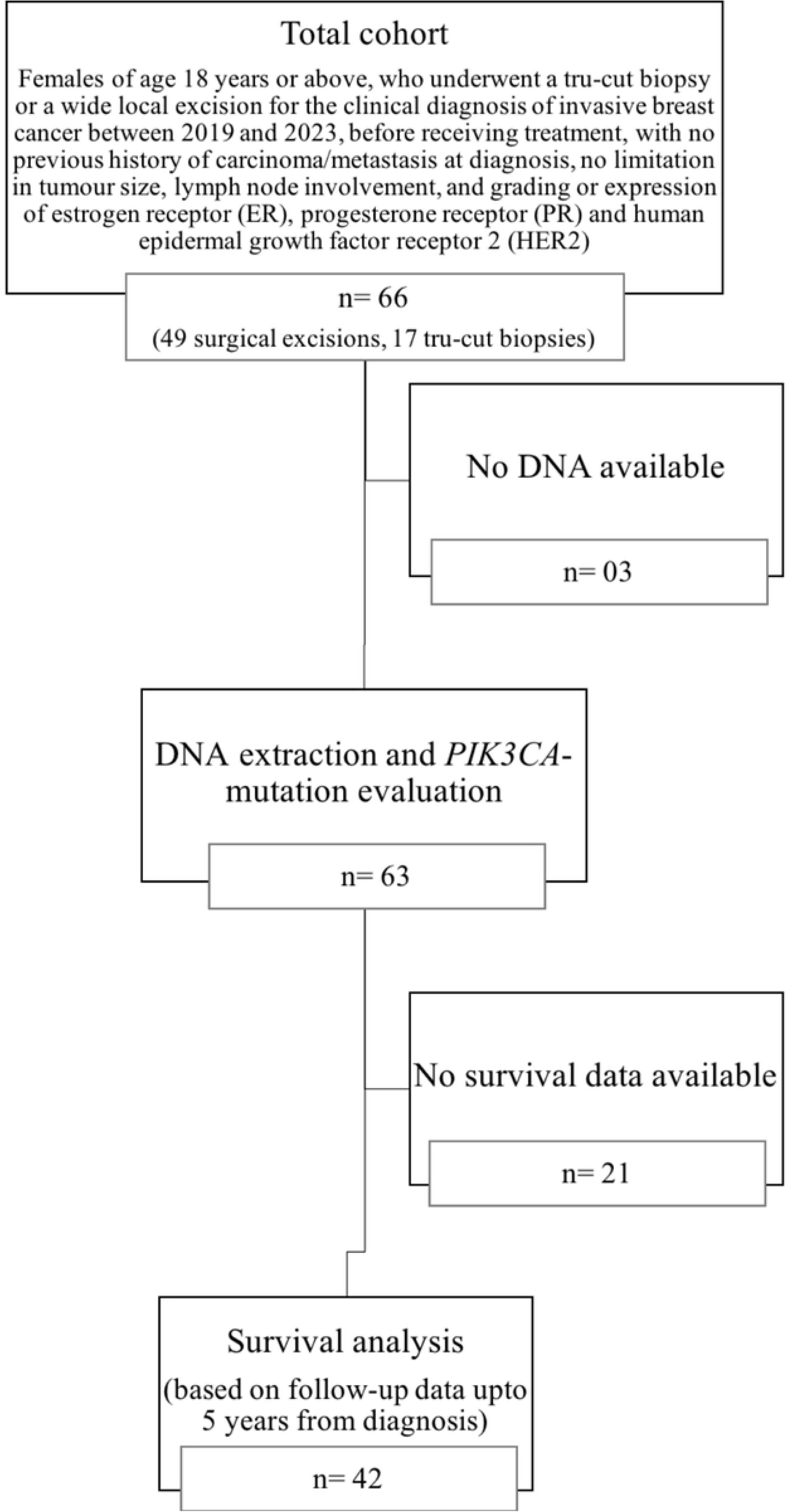
Flowchart of patient enrollment, inclusion criteria, and study workflow.

To ensure patient anonymity, medical and follow-up data of individual patients were recorded, and the samples were identified exclusively using identification numbers assigned to each sample for the purpose of research. Therefore, as authors did not have access to information that could identify individual participants during or after data collection, total anonymity of patient identity was maintained throughout the study.

### DNA extraction and hotspot mutation assay

Genomic DNA was extracted from FFPE samples using the QIAamp DNA FFPE Tissue Kit according to the manufacturer’s instructions (Cat. No. 56404; Qiagen, Hilden, Germany) and assessed for quality and quantity using the Thermo Scientific^TM^ NanoDrop^TM^ One Spectrophotometer. The three hotspot mutations of the *PIK3CA* oncogene-H1047R, E545K, and E542K-were genotyped in 63 of the selected samples using the QClamp® *PIK3CA* Mutation Detection Test Kit (Cat. No. DC-10-1072R; DiaCarta, CA 94588, USA), which utilized a Xeno-nucleic acid (XNA)-based mutation-specific quantitative real-time polymerase chain reaction (qPCR) method.

### Endpoint and statistical analysis

The prevalence of *PIK3CA* mutations in the study cohort and their associations with clinicopathological features were determined using the χ2 test. Fisher’s exact test was employed for dichotomous features. To enhance the statistical validity and minimize the risk of type I errors, patients were grouped according to significant, widely accepted clinicopathological cutoffs in oncology based on the sample size available for each category (S1 Table). In the analysis of ancestry, patients grouped into Indo-Aryan ancestry (Sinhalese & Burghers) were compared to those of Dravidian ancestry (Tamils & Moors). Depending on data normality, either a t-test or Mann-Whitney Test was performed to compare the mean or median of populations for the relevant variables. For specific variables showing significant associations, binary logistic regression was used to obtain the odds ratio (OR) and the 95% confidence interval (CI). This binary logistic regression model assumed that there were no significant multicollinearity issues, and all independent variables were linearly related to the log-odds of the dependent variable.

All patients underwent routine clinical examinations at the hospital every 3-6 months, and the clinicopathological variables analyzed were those recorded at the initial diagnosis. Since all patients with survival data (n=42) were alive at the last follow-up, RFS was used as the survival metric instead of OS, and was defined as the time from the date of curative surgery to the clinical diagnosis of recurrence. Events considered in RFS included distant and local invasive recurrence. Patients without survival data were excluded from the survival analysis. The median follow-up time after diagnosis was 37.00 months (0.00–81.19).

The Kaplan-Meier method was used to plot RFS, while the Mantel-Cox log-rank test was applied to compare different subgroups of the variables. Significant parameters identified by the log-rank test (p<0.05) were further analyzed using a univariate Cox proportional hazard regression model. A multivariate model was finally used to analyze the combined effect of all significant associations on RFS. However, for parameters which could not generate a statistically robust Cox proportional hazard regression model due to limited data on survival, only the p-value was reported.

Samples with missing data for the variable of interest were excluded from specific analyses. All statistical tests were two-sided and a p-value below 0.05 was considered statistically significant. Analyses were performed using SPSS 25 (Released 2017. IBM SPSS Statistics for Windows, Version 25.0. Armonk, NY: IBM Corp.) and R (version 4.0.2, R Foundation for Statistical Computing, Vienna, Austria).

## Results

### Analysis of the clinicopathological features and ethnicity

The studied cohort consisted of 66 patients clinically diagnosed with breast cancer between 2019 and 2023. The median age at the time of diagnosis was 58.00 years (S2 Fig).

Table 1 presents several key characteristics of the cohort. Accordingly, IHC analysis revealed that over 80% of the tumors were SHR-positive HER2-negative luminal subtypes, with luminal A and B subtypes occurring in equal proportions. Over 95% of the cohort comprised of Sinhalese patients while the remaining 5% included two Tamil patients, one Moor, and one Burgher patient. This represents only a rough approximation of the actual ethnic distribution in Sri Lanka, where approximately 72% of the population are Sinhalese [23].

**Table 1.**
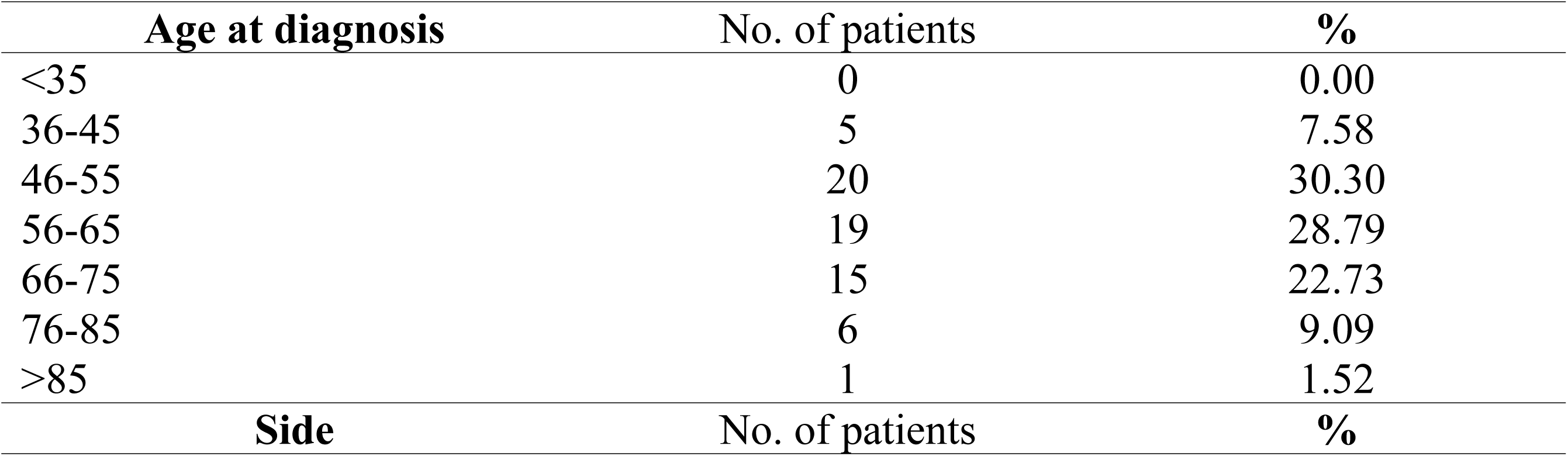
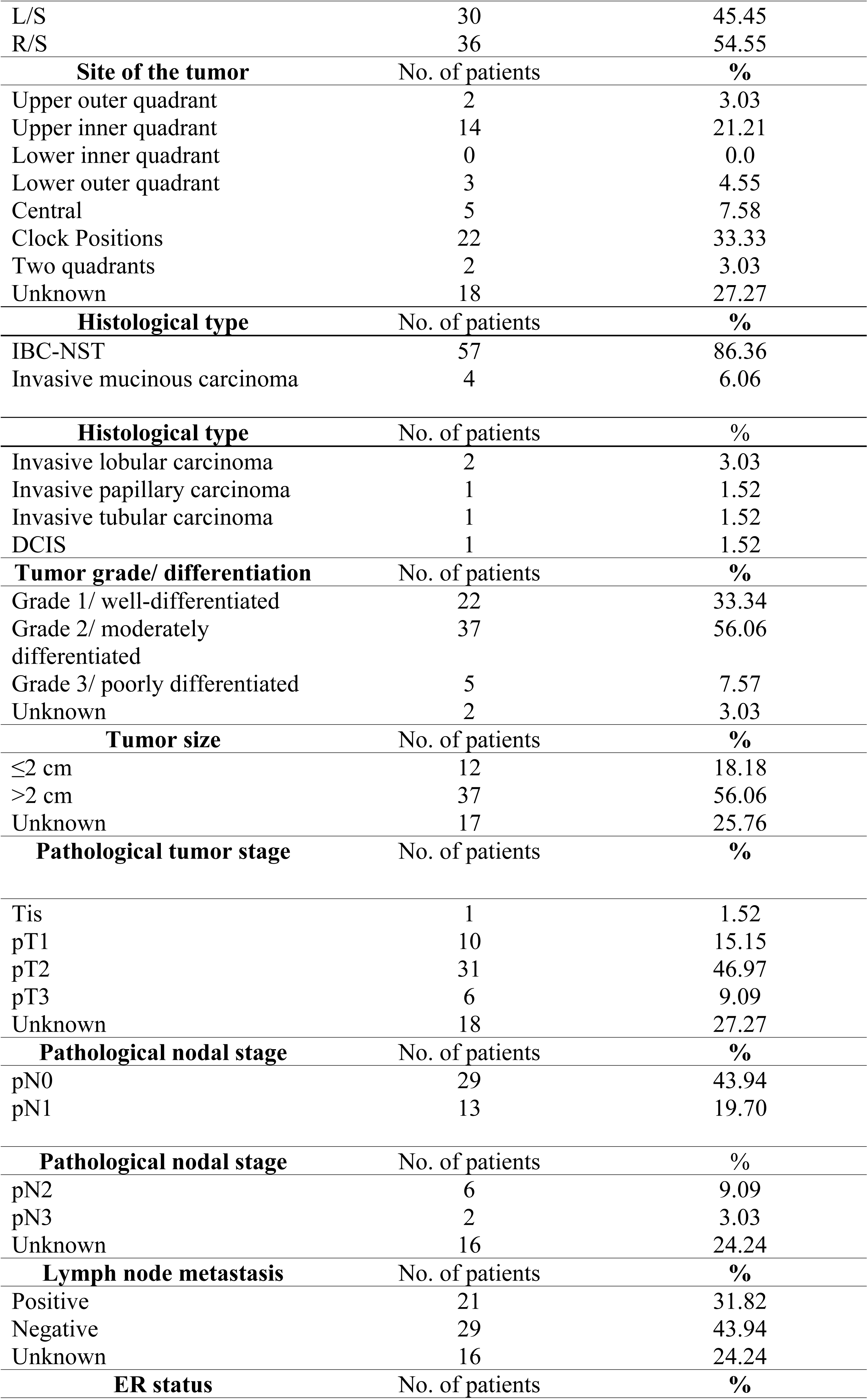
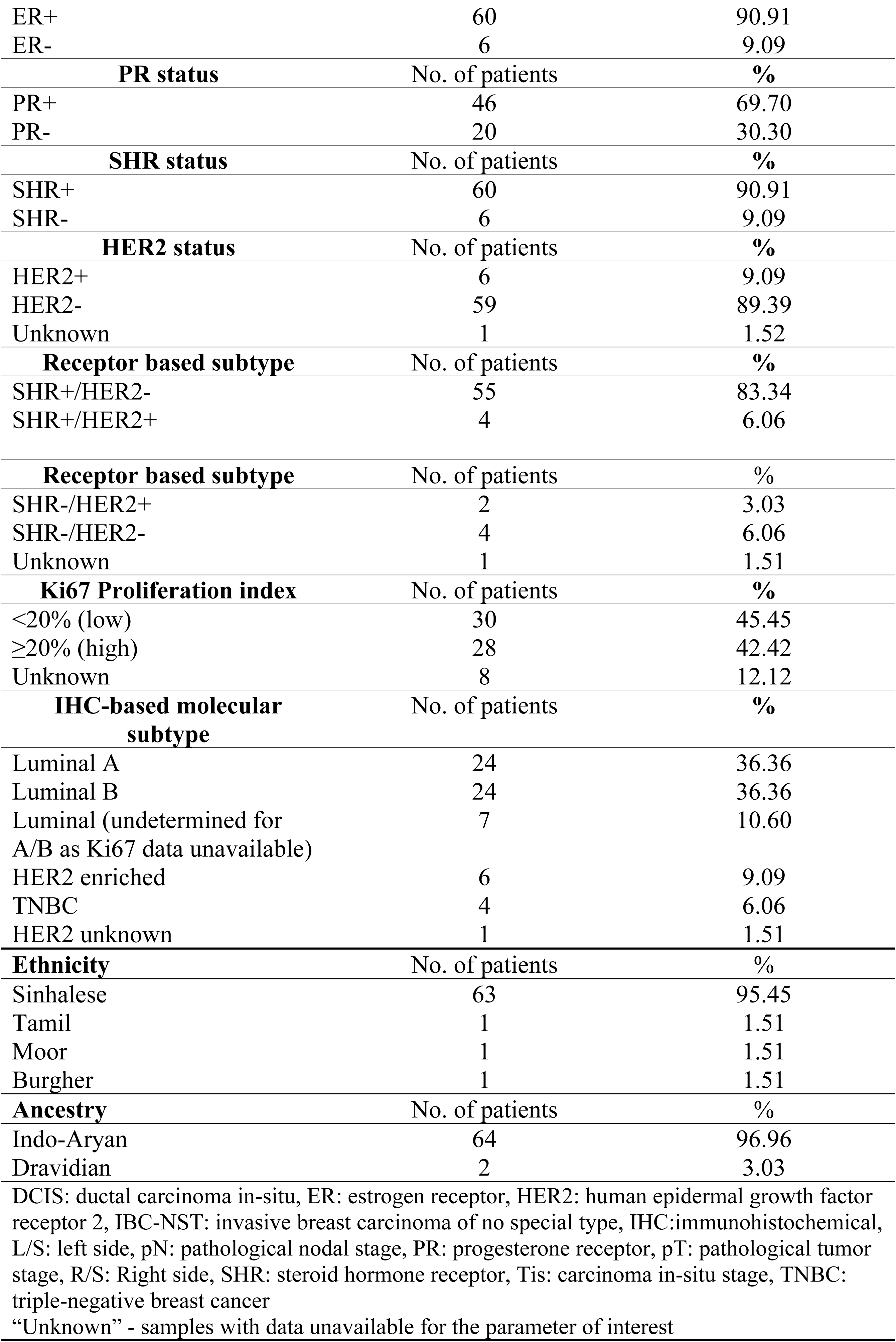
Clinicopathological and population characteristics of the cohort.

| Age at diagnosis | No. of patients | % |
| --- | --- | --- |
| <35 | 0 | 0.00 |
| 36-45 | 5 | 7.58 |
| 46-55 | 20 | 30.30 |
| 56-65 | 19 | 28.79 |
| 66-75 | 15 | 22.73 |
| 76-85 | 6 | 9.09 |
| >85 | 1 | 1.52 |
| Side | No. of patients | % |
| L/S | 30 | 45.45 |
| R/S | 36 | 54.55 |
| **Site of the tumor** | **No. of patients** | **%** |
| Upper outer quadrant | 2 | 3.03 |
| Upper inner quadrant | 14 | 21.21 |
| Lower inner quadrant | 0 | 0.0 |
| Lower outer quadrant | 3 | 4.55 |
| Central | 5 | 7.58 |
| Clock Positions | 22 | 33.33 |
| Two quadrants | 2 | 3.03 |
| Unknown | 18 | 27.27 |
| **Histological type** | **No. of patients** | **%** |
| IBC-NST | 57 | 86.36 |
| Invasive mucinous carcinoma | 4 | 6.06 |
| **Histological type** | **No. of patients** | **%** |
| Invasive lobular carcinoma | 2 | 3.03 |
| Invasive papillary carcinoma | 1 | 1.52 |
| Invasive tubular carcinoma | 1 | 1.52 |
| DCIS | 1 | 1.52 |
| **Tumor grade/ differentiation** | **No. of patients** | **%** |
| Grade 1/ well-differentiated | 22 | 33.34 |
| Grade 2/ moderately differentiated | 37 | 56.06 |
| Grade 3/ poorly differentiated | 5 | 7.57 |
| Unknown | 2 | 3.03 |
| **Tumor size** | **No. of patients** | **%** |
| ≤2 cm | 12 | 18.18 |
| >2 cm | 37 | 56.06 |
| Unknown | 17 | 25.76 |
| **Pathological tumor stage** | **No. of patients** | **%** |
| Tis | 1 | 1.52 |
| pT1 | 10 | 15.15 |
| pT2 | 31 | 46.97 |
| pT3 | 6 | 9.09 |
| Unknown | 18 | 27.27 |
| **Pathological nodal stage** | **No. of patients** | **%** |
| pN0 | 29 | 43.94 |
| pN1 | 13 | 19.70 |
| **Pathological nodal stage** | **No. of patients** | **%** |
| pN2 | 6 | 9.09 |
| pN3 | 2 | 3.03 |
| Unknown | 16 | 24.24 |
| **Lymph node metastasis** | **No. of patients** | **%** |
| Positive | 21 | 31.82 |
| Negative | 29 | 43.94 |
| Unknown | 16 | 24.24 |
| **ER status** | **No. of patients** | **%** |
| ER+ | 60 | 90.91 |
| ER- | 6 | 9.09 |
| **PR status** | No. of patients | % |
| PR+ | 46 | 69.70 |
| PR- | 20 | 30.30 |
| **SHR status** | No. of patients | % |
| SHR+ | 60 | 90.91 |
| SHR- | 6 | 9.09 |
| **HER2 status** | No. of patients | % |
| HER2+ | 6 | 9.09 |
| HER2- | 59 | 89.39 |
| Unknown | 1 | 1.52 |
| **Receptor based subtype** | No. of patients | % |
| SHR+/HER2- | 55 | 83.34 |
| SHR+/HER2+ | 4 | 6.06 |
| **Receptor based subtype** | No. of patients | % |
| SHR-/HER2+ | 2 | 3.03 |
| SHR-/HER2- | 4 | 6.06 |
| Unknown | 1 | 1.51 |
| **Ki67 Proliferation index** | No. of patients | % |
| <20% (low) | 30 | 45.45 |
| ≥20% (high) | 28 | 42.42 |
| Unknown | 8 | 12.12 |
| **IHC-based molecular subtype** | No. of patients | % |
| Luminal A | 24 | 36.36 |
| Luminal B | 24 | 36.36 |
| Luminal (undetermined for A/B as Ki67 data unavailable) | 7 | 10.60 |
| HER2 enriched | 6 | 9.09 |
| TNBC | 4 | 6.06 |
| HER2 unknown | 1 | 1.51 |
| **Ethnicity** | No. of patients | % |
| Sinhalese | 63 | 95.45 |
| Tamil | 1 | 1.51 |
| Moor | 1 | 1.51 |
| Burgher | 1 | 1.51 |
| **Ancestry** | No. of patients | % |
| Indo-Aryan | 64 | 96.96 |
| Dravidian | 2 | 3.03 |

DCIS: ductal carcinoma in-situ, ER: estrogen receptor, HER2: human epidermal growth factor receptor 2, IBC-NST: invasive breast carcinoma of no special type, IHC:immunohistochemical, L/S: left side, pN: pathological nodal stage, PR: progesterone receptor, pT: pathological tumor stage, R/S: Right side, SHR: steroid hormone receptor, Tis: carcinoma in-situ stage, TNBC: triple-negative breast cancer “Unknown” - samples with data unavailable for the parameter of interest

### Prevalence of *PIK3CA* mutations in the cohort

Missense mutations in the *PIK3CA* oncogene were detected in 17.46% (n=11) of the cohort. The mutations observed included H1047R (c.3140A>G) in 81.81% (n=9) and E542K (c.1624G>A) in 18.18% (n=2) of the mutated samples. Notably, both H1047R and E542K mutations were detected in one sample (co-mutation prevalence: 9.09% of the mutated and 1.58% of the total samples). No E545K mutations (c.1633G>A) were detected in the cohort.

### Association of *PIK3CA* mutations with clinicopathological parameters and ancestry

All *PIK3CA* mutations identified were found in SHR+/HER2-luminal tumors (Table 2). Importantly, a significant association was observed between *PIK3CA* mutational status and lymph node (LN) metastasis, with patients harboring a *PIK3CA* mutation exhibiting 9.60 times higher odds of LN metastasis compared to those without a *PIK3CA* mutation (p=0.036, OR 9.60, 95% CI 1.05-87.78). Notably, a significant association was observed between the ancestry of the patients and the presence of a *PIK3CA* mutation (p=0.028). Other parameters showed no significant association with the mutational status.

**Table 2.**
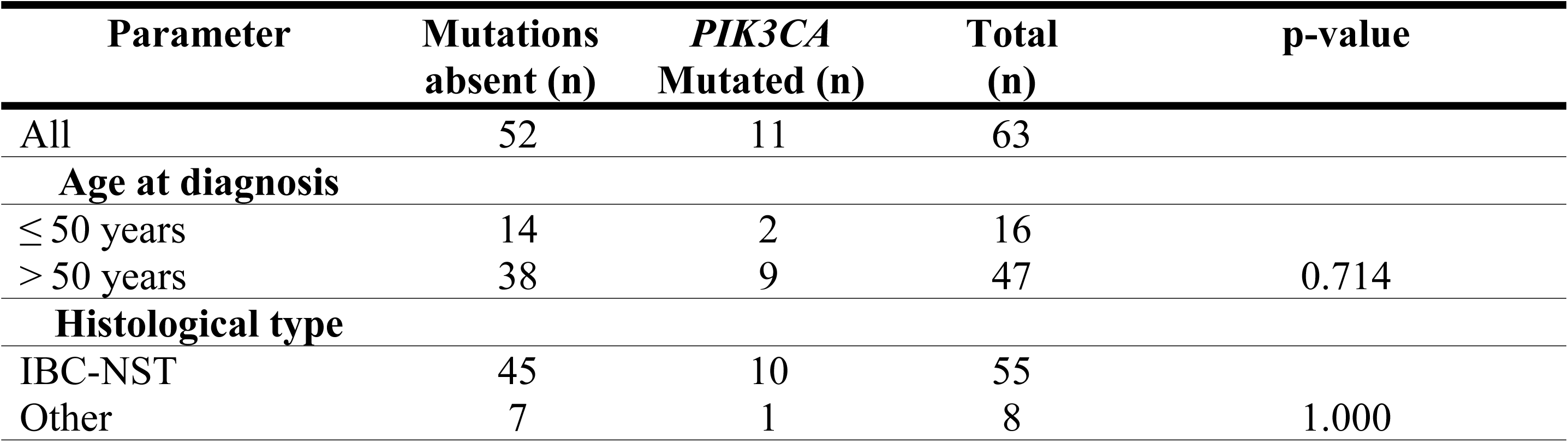
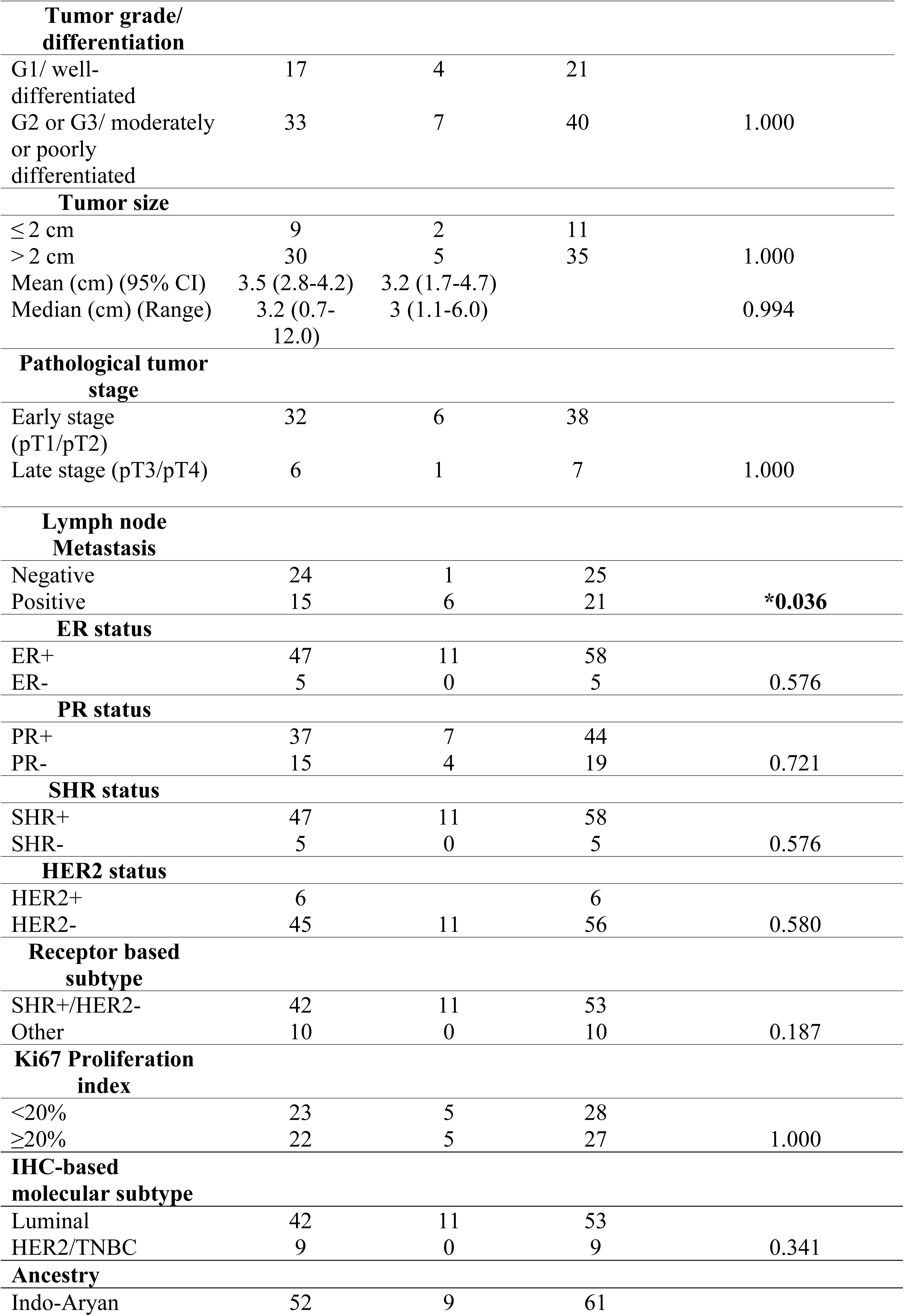
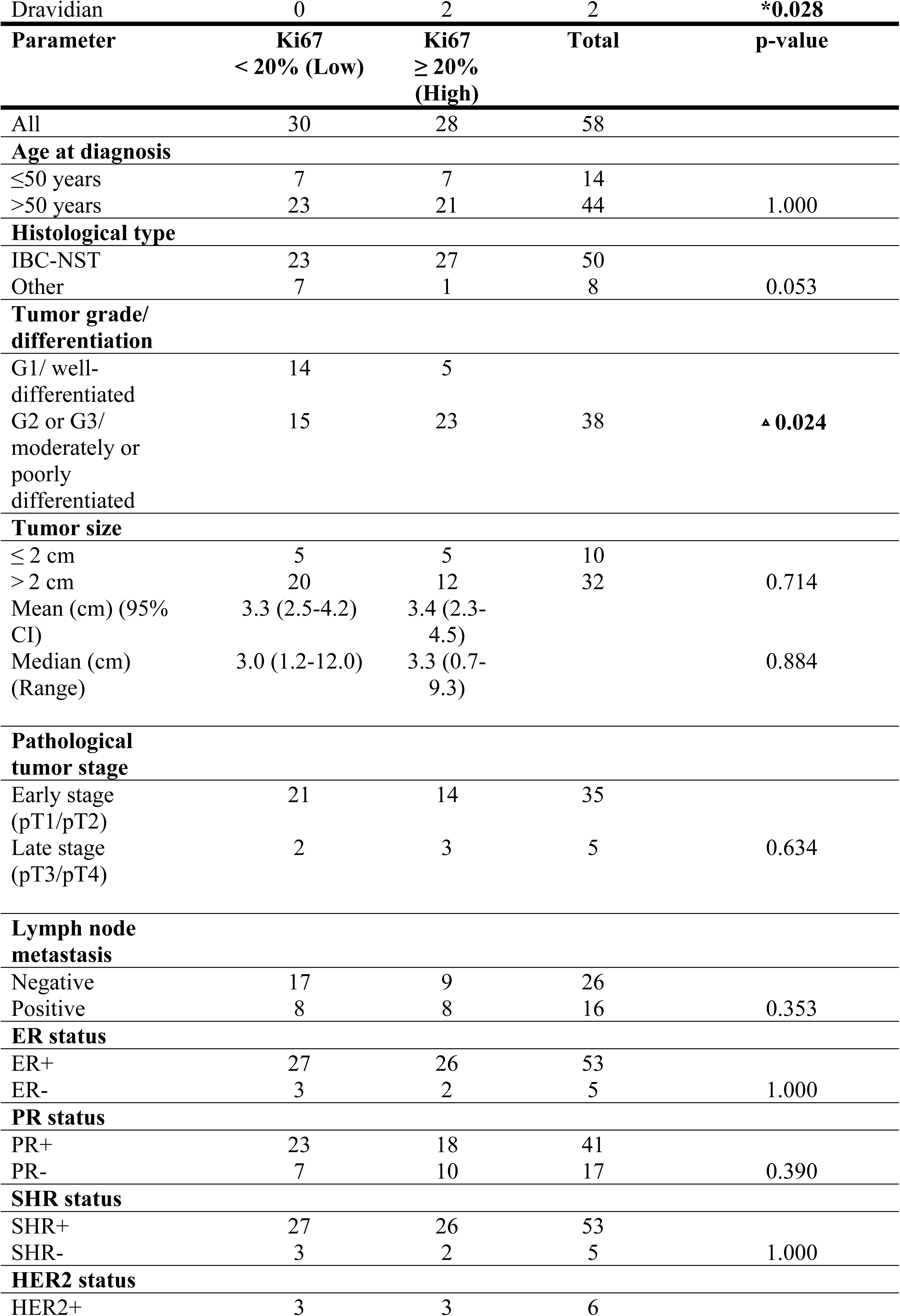
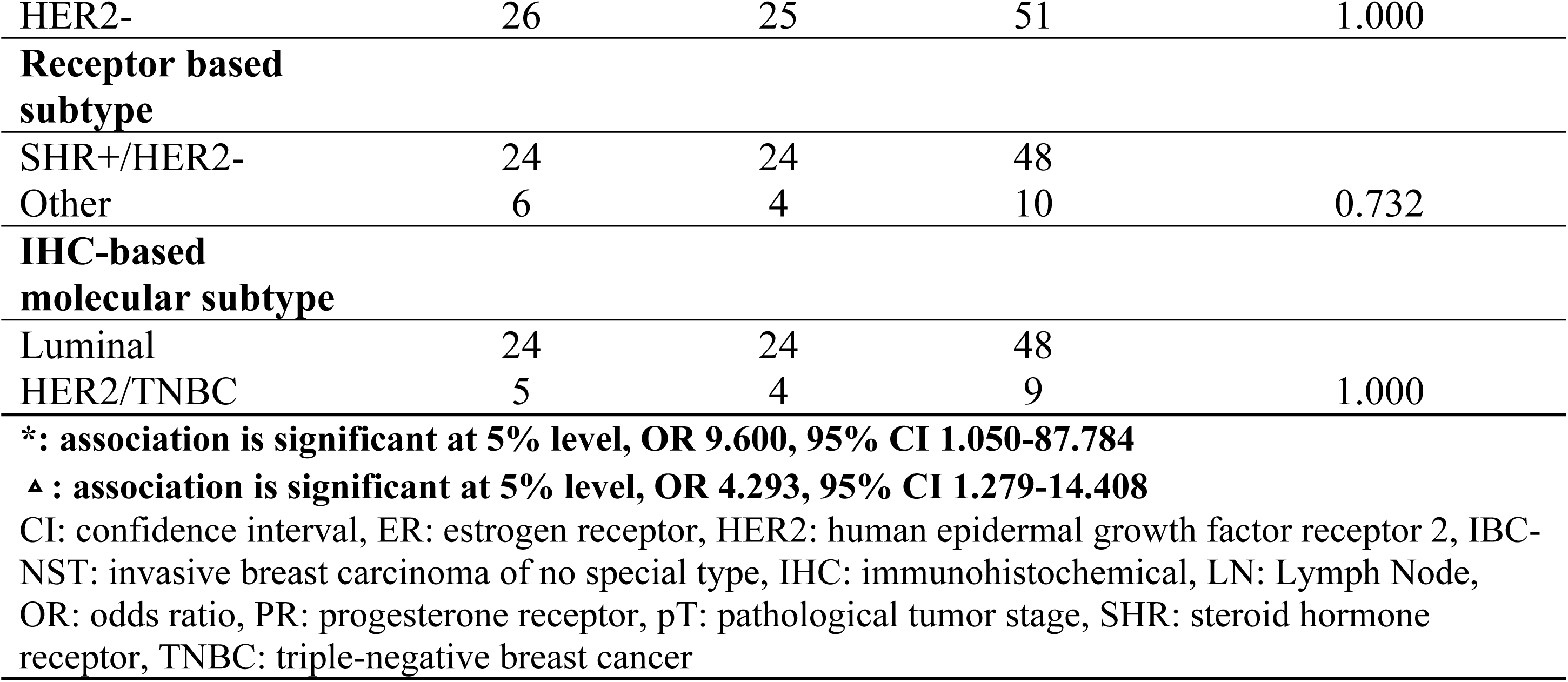
*PIK3CA* mutational status and Ki67 proliferative index in selected clinicopathological and ancestral groups.

### Associations among other clinical and histopathological parameters

Table 2 also outlines the associations between the Ki67 proliferation index and other clinicopathological features. A significant association was identified between Ki67 status and tumor grade, with higher Ki67 proliferative indices being linked to a 4.29 times higher likelihood of developing moderately or poorly differentiated breast cancers of higher tumor grades, compared to those with proliferative indices < 20% (p=0.024, OR 4.29, 95% CI 1.28-14.41).

No statistically significant associations were observed between LN metastasis or patient age group and other clinicopathological parameters (S2 Table).

### Impact of *PIK3CA* mutational status on RFS

Among patients with survival data, 9.5% (n=4) experienced a relapse in a different organ (e.g., pelvic bone and lungs) following curative surgery. Three of these patients (75.0%) had *PIK3CA* mutations, with two harboring the H1047R mutation (50%) and one carrying the E542K mutation (25%).

The presence of a *PIK3CA* mutation was thereby significantly associated with reduced RFS (p<0.001), as shown in the Kaplan-Meier curve (Fig 2A and Table 3). Patients with a *PIK3CA* mutation had 26.19 times higher odds of relapse compared to the those without a mutation (HR 26.19, 95% CI 2.67-256.47). The mean RFS for patients with *PIK3CA* mutations was 26.5 months.

**Fig 2.**
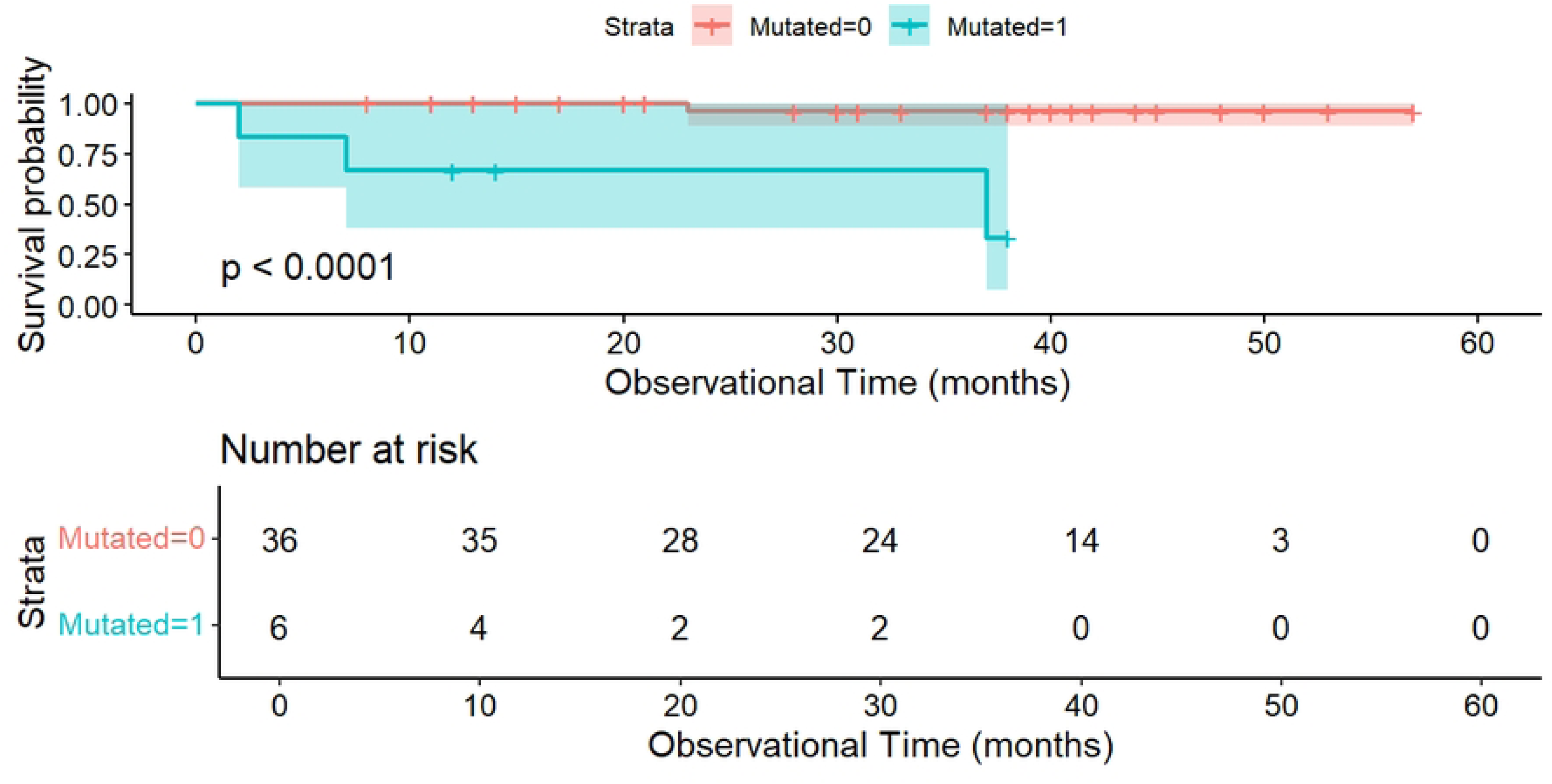
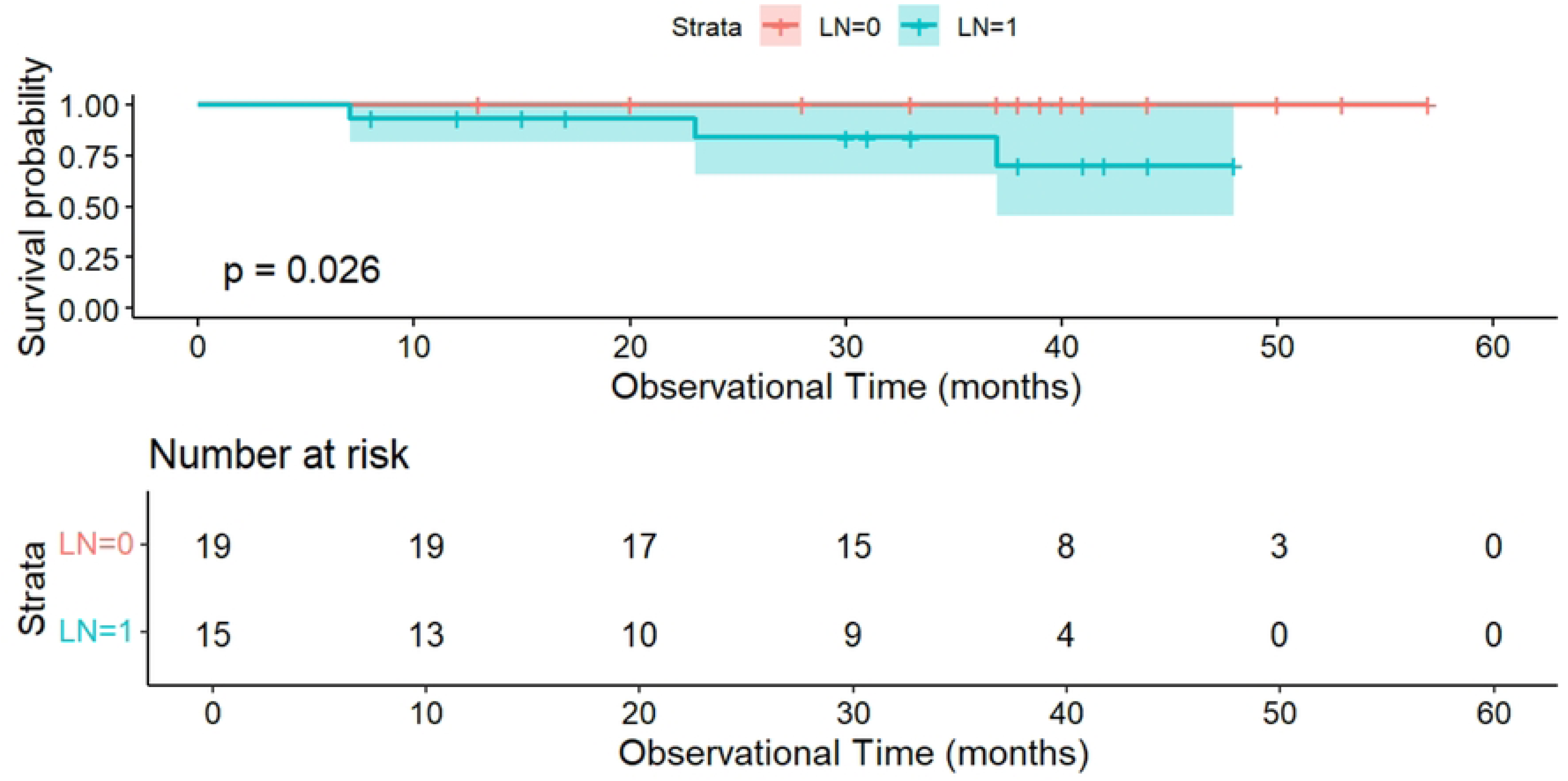
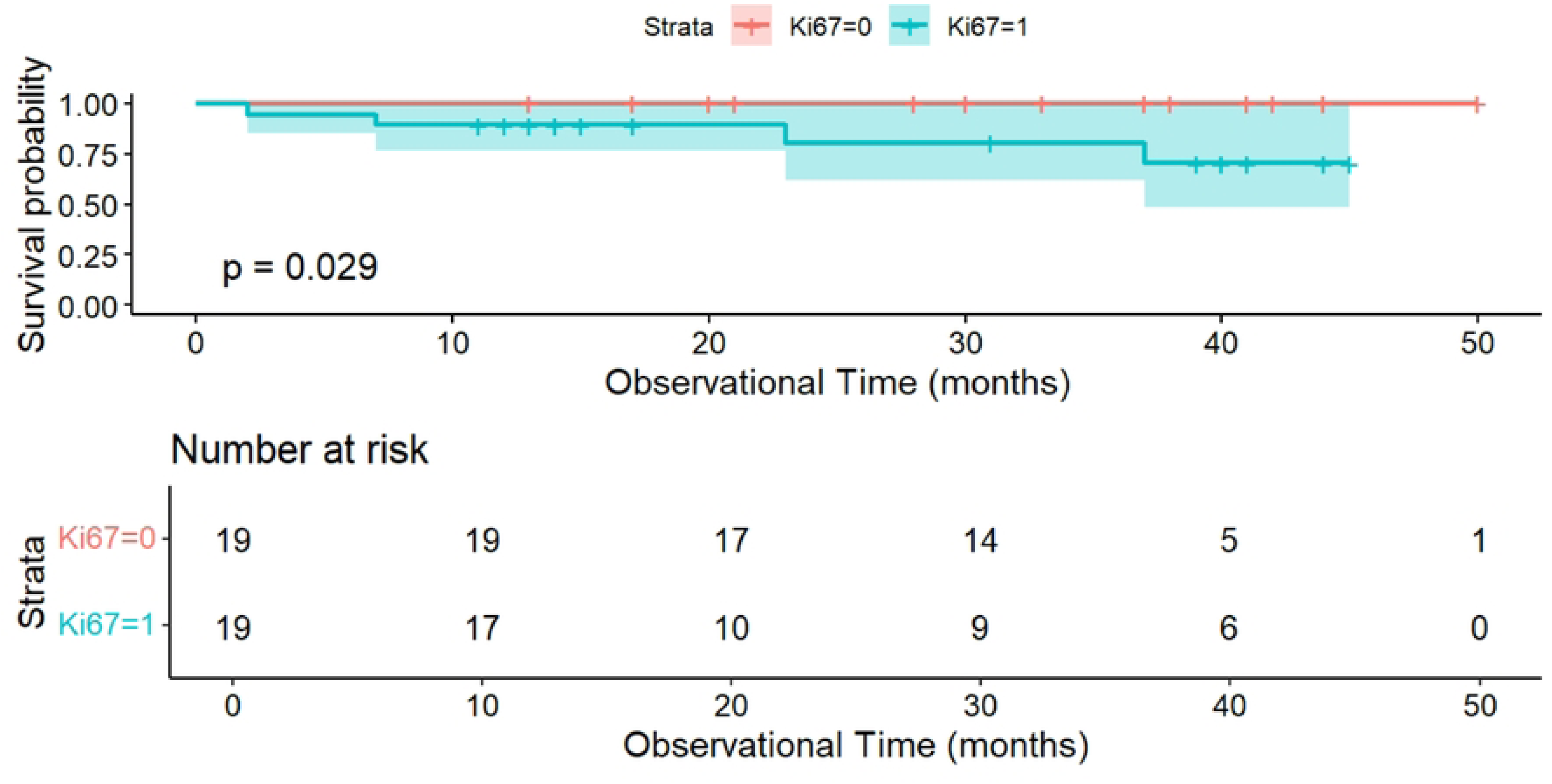
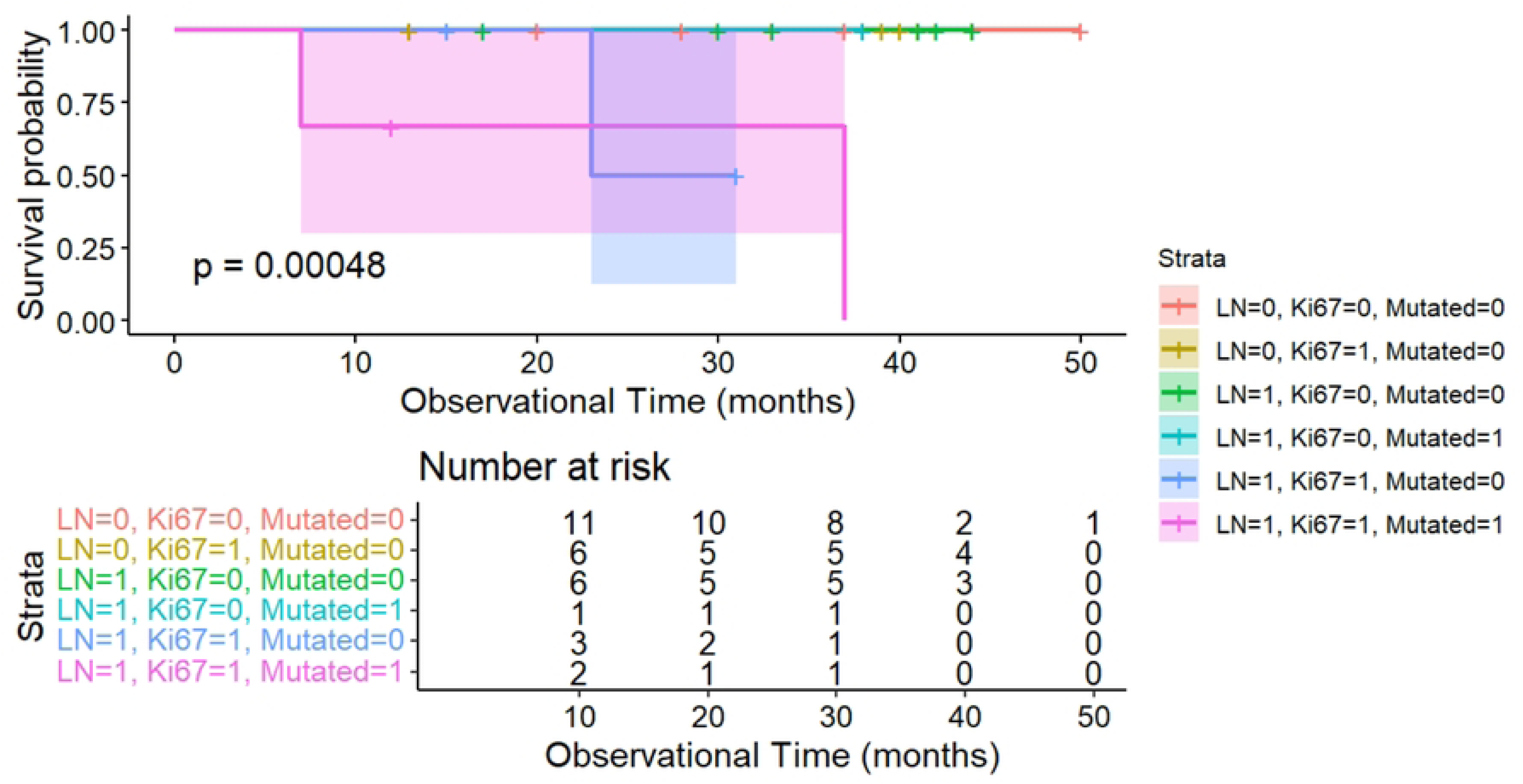
Kaplan-Meier survival estimates for relapse-free survival (RFS) stratified by *PIK3CA* mutation status (A), status of lymph node (LN) metastasis (B) and Ki67 proliferative status (C) while (D) shows the multivariate impact of all above three factors – *PIK3CA* mutation, LN metastasis and Ki67 index on RFS. Fig. 2A: p-value<0.001, HR 26.187, 95% CI 2.674 – 256.475 Fig. 2B: p-value=0.026, HR 123.939, 95% CI could not be generated with statistical robustness due to small sample size Fig. 2C: p-value=0.029, HR 79.688, 95% CI could not be generated with statistical robustness due to small sample size Fig.2D: p-value=0.00048, HR and 95% CI could not be generated with statistical robustness due to small sample size HR: hazard ratio, CI: confidence interval

**Table 3.**
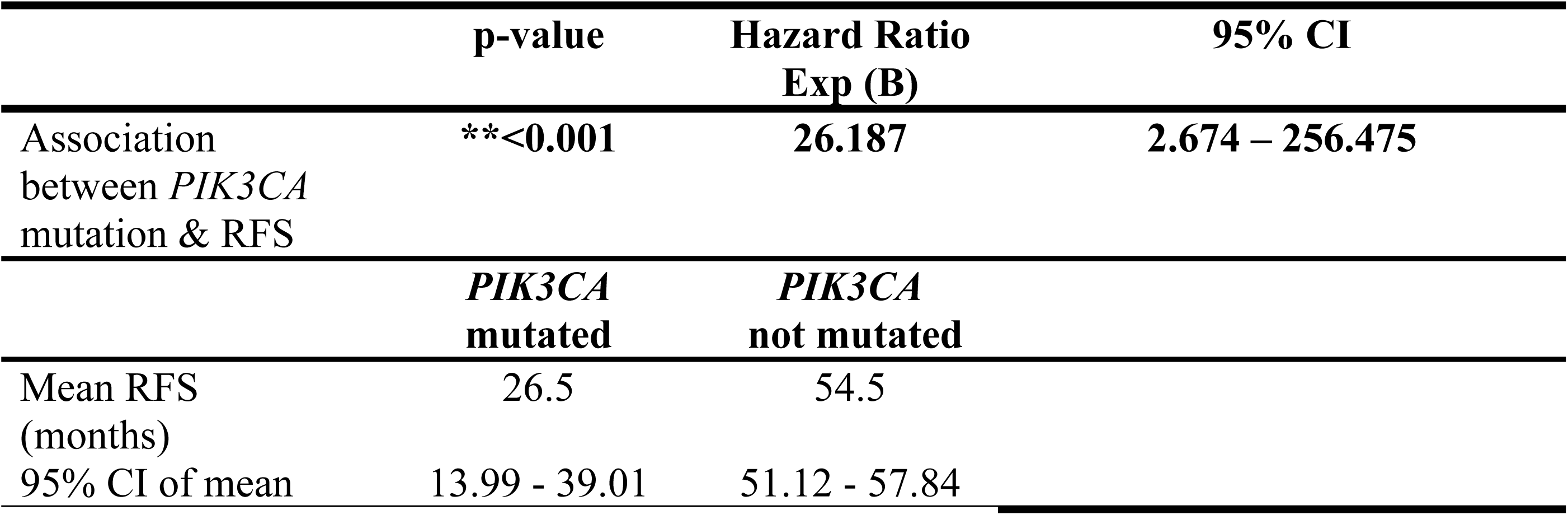
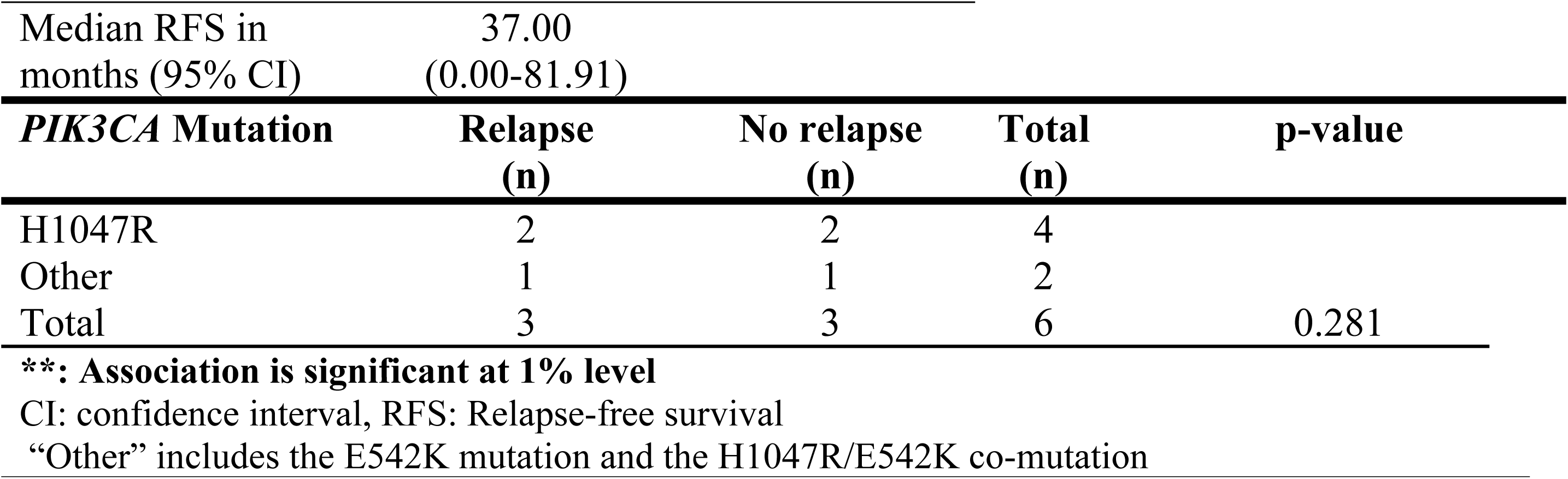
Impact of the overall and specific *PIK3CA* mutation status on relapse-free survival (RFS)

However, no significant association was found between individual *PIK3CA* hotspot mutations (H1047R or E542K) and RFS (Table 3).

### Impact of selected clinicopathological features on RFS

LN metastasis was significantly associated with RFS (Table 4). Patients with LN-positive status had 123.94 times higher hazard of relapse compared to LN-negative patients (p=0.026, HR 123.94; Fig 2B). Additionally, a significant association was observed between Ki67 proliferative status and RFS, with patients having higher proliferative indices facing a 79.69 times higher hazard of relapse compared to those with Ki67 indices <20% (p=0.029, HR 79.69; Fig 2C).

**Table 4.**
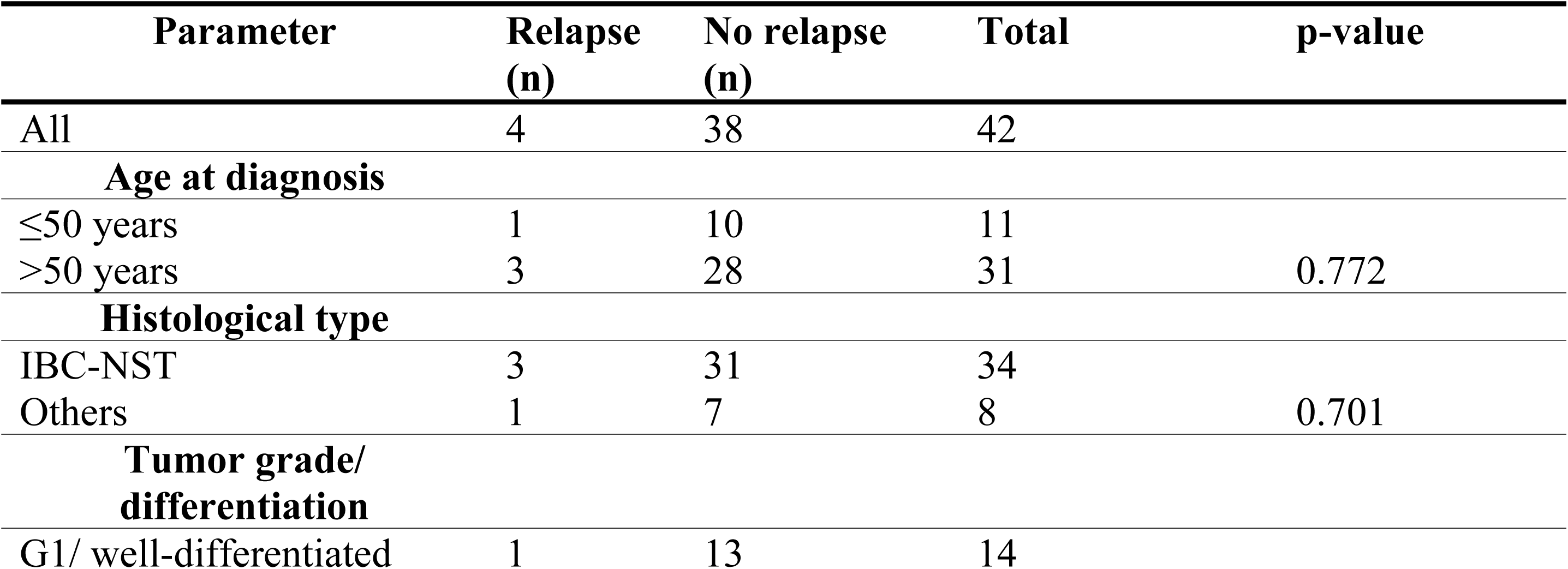
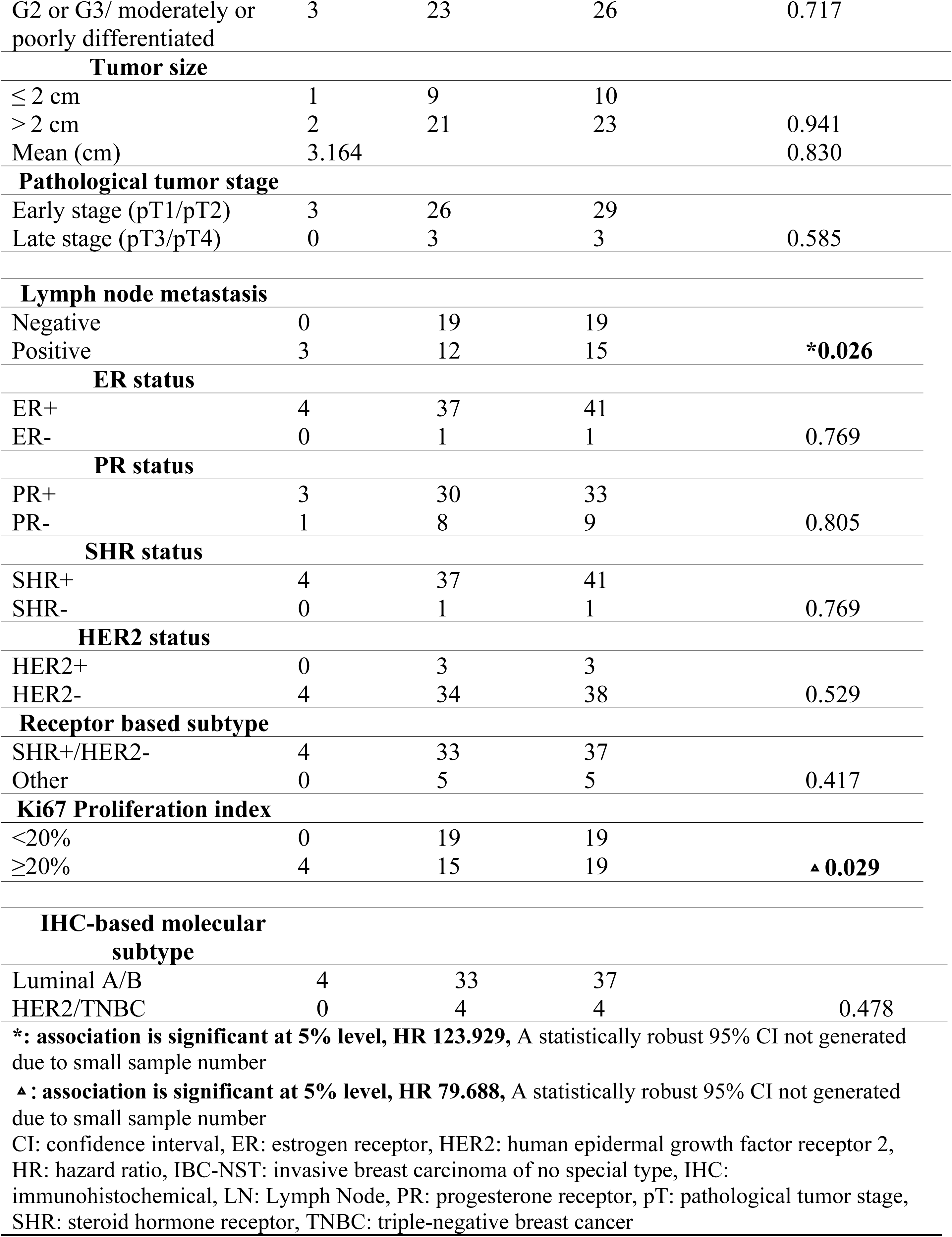
Univariate analysis of relapse-free survival (RFS) with selected clinicopathological parameters.

No other clinicopathological characteristics showed significant associations with RFS.

Finally, the multivariate Cox proportional hazard model (Fig 2D) demonstrated a significant association between the presence of a *PIK3CA* mutation, LN metastasis and a high Ki67 index with RFS (p<0.001).

## Discussion

This study, to our knowledge, is the first to profile *PIK3CA* mutations, assess their prognostic value, and evaluate the implications of patient ancestry on somatic mutations in Sri Lankan breast cancer patients. Despite being a pilot study conducted in an underrepresented island population, our findings lay the groundwork for future research, including meta-analyses in larger cohorts.

The analysis of the clinicopathological parameters showed a median age of 58 years at breast cancer diagnosis, higher than the median age reported in a Sri Lankan study done over a decade ago, and higher than those reported from several Southeast Asian countries and Arab countries [26–29]. This could be due to Sri Lanka having one of the fastest ageing populations in the world [26]. The exponentially growing population of older women [26] would thus raise the incidence of breast cancer among this demographic, making it a key factor in the observed age shift in diagnosis. However, the fact that the majority of patients were already at stage 2 at the time of diagnosis, even with slow-growing cancers (i.e., luminal A and B), suggests a much earlier onset of the disease. This trend, consistent with other Asian countries, could probably be attributed to similar genetic and sociocultural factors and contrasts with developed Western countries, where median age of diagnosis occurs much later [30].

Similar to reports by previous studies conducted in Sri Lanka [31,32], it is noticeable that a majority of the tumors in our cohort were invasive breast carcinomas (IBC-NST) of tumor grade 2 and/or stage II at diagnosis. This could be likely due to the absence of an established screening program aimed at early detection in Sri Lanka, thus reducing the likelihood of the cancer being detected at stage I and worsening disease outcomes [31]. These observations therefore highlight the need for the implementation of an effective screening system for breast cancer in Sri Lanka, to facilitate early diagnosis and ultimately reduce the overall burden of breast cancer in the country. Consistent with both local and global statistics [31–33], most tumors were ER/PR+ and HER2-, confirming that SHR+/HER2-luminal subtypes are the most common in Sri Lanka.

One of the study’s primary objectives was to catalog *PIK3CA* mutations in the cohort of patients and to detect associations with clinicopathological features. The detection of H1047R and E542K mutations in 17.46% of cases falls within the prevalence range of *PIK3CA* mutations reported in many global studies on breast cancer [5–10]. Notably, this study also provides the first evidence of *PIK3CA* mutations in Sri Lankan breast cancer patients. India, the closest nation to Sri Lanka both geographically and genetically, reports a *PIK3CA* mutation prevalence of 23.2% [34], closely aligned with our findings. This similarity likely reflects common hereditary factors shaped by historical migrations and cultural exchanges, which may have influenced the prevalence of *PIK3CA* mutations in the South Asian region.

Although the predominance of H1047R as the most frequently observed *PIK3CA* mutation mirrors observations in other populations [35], our study reports a much a higher frequency of 81.81% for H1047R among the total *PIK3CA* mutations, compared to the frequency (≈58%) reported in other studies [35,36]. A particularly rare and unexpected observation was the absence of the E545K mutation in our cohort, which is commonly reported as a moderately prevalent *PIK3CA* mutation (≈10%) in many other global populations [37,38]. This observation aligns with findings from an Iranian study, where the E545K mutation was similarly absent [39]. Given that over 96% of our cohort comprises Sri Lankan Sinhalese and Sri Lankan Burghers (both of Indo-Aryan ancestry), this shared absence is likely to be stemming from a common ancestral link with Iranians, as both Iranians and Indo-Aryans are descendants of the Indo-Iranian branch of the Indo-European language family [23,24,40]. The absence of the E545K mutation, may thus be tied to ancestral roots, and consequent genetic backgrounds, among other possible factors. This provides valuable preliminary data from Sri Lanka, contributing to the global effort of cataloging *PIK3CA* mutations, which are highly heterogeneous across populations with different ancestral origins [10].

The significant association between patient ancestry and the presence of a *PIK3CA* mutation (p=0.028), where the Indo-Aryan and Dravidian ancestries could potentially be associated with differing rates of *PIK3CA* mutations, suggests that ancestral influence tied to ethnic backgrounds could affect the occurrence of *PIK3CA* mutations in Sri Lanka as well as in South Asia. Though the strength of this observation is hampered by the small sample size, particularly with respect to the patients of Dravidian origin (one Tamil and one Moor in the entire cohort), the presence of *PIK3CA* mutations in both of them cannot be disregarded and warrant further ancestry-oriented mutational analyses. While this association aligns with global findings that somatic *PIK3CA* mutations are linked with ancestry across all cancer types including breast cancer [19–22], our study fills an important gap by investigating these ancestry-specific associations in a cohort, exclusively of South Asian ancestry. These insights may have significant implications for personalized treatment strategies, which may need to be tailored based on the patient’s ancestry, as mutation profiles observed in one ancestry may not apply to others – in turn influencing treatment efficacy and outcomes.

Another notable observation was that the *PIK3CA* mutations in the cohort were exclusively seen in SHR+/HER2-luminal tumors, consistent with a similar subtype-specific dominance of the mutation in previous studies [9]. The molecular mechanisms, including cross talk between the PI3K/AKT/mTOR and ER signaling pathways are postulated to be mediating this, with *PIK3CA* mutations driving oncogenesis via ER signaling, thereby predominating in SHR+ breast cancer subtypes [17].

The significant association between *PIK3CA* mutations and increased likelihood of axillary LN metastasis (p=0.036, OR 9.60) highlights the potential prognostic value of *PIK3CA* mutations in predicting breast cancer progression in the Sri Lankan context. LN metastasis typically indicates a more advanced disease stage, a higher risk of distant metastases and worse outcomes [41]. Therefore, *PIK3CA* mutation testing could guide therapeutic strategies, particularly in decisions related to axillary management and risk assessment, with more intensive therapeutic approaches needed by patients with tumor-positive axillary LNs [42]. Divergent findings in other studies [43], may reflect differences in genetic makeup and lifestyle factors, further emphasizing the need for nuanced molecular investigations in underrepresented populations like Sri Lanka.

The absence of associations between clinicopathological features and age group may indicate that other parameters, such as histological grade and Ki67 index, hold greater prognostic value than age, particularly in early-stage breast cancer cohorts [44], like ours. This is supported by the significant association between higher Ki67 indices and an increased risk of developing higher-grade tumors (p=0.024, OR 4.29). These findings further support the use of Ki67 index as a valuable prognostic marker [45] in Sri Lankan breast cancer patients and its importance in clinical decision-making.

The striking association between *PIK3CA* mutations and reduced RFS in Sri Lankan breast cancer patients, with high statistical significance (p<0.001, HR 26.19, 95% CI 2.67 – 256.47) suggests that these mutations have a considerable impact on disease prognosis. The broad confidence interval reflects the small sample size, but even the lower bound presents a significantly increased risk of relapse, nearly three times greater than patients without a *PIK3CA* mutation. This implies a significant impact of *PIK3CA* mutations on disease progression and brings out its potential clinical relevance as a prognostic biomarker in Sri Lankan breast cancer patients. As the first study to analyze *PIK3CA* mutations in Sri Lankan breast cancer patients, our findings are consistent with results from other Asian studies as well as studies on other malignancies, such as colorectal cancer [13,46,47].

However, our results did not show mutation-specific associations with RFS, which may be attributed to the smaller cohort size, necessitating further investigation with larger studies. Nevertheless, the predominance of the H1047R mutation-with an observed frequency of 0.81 within the total mutations, suggests that it is likely to be responsible for reducing RFS [35] in our cohort, mirroring findings of the aforementioned Iranian study [39]. Given the shared Indo-Iranian ancestry of the Sinhalese and Iranians, this parallel may hint at ancestral genetic traits influencing both mutation occurrence and its clinical impact. This further emphasizes the need for nuanced investigations and personalized treatment approaches in populations with specific ancestral backgrounds.

Our analysis of clinicopathological characteristics also revealed that LN metastasis and Ki67 proliferative index are key determinants of relapse risk. Patients with either tumor-positive LNs or a higher Ki67 index (≥20%), exhibited a significantly greater hazard of relapse compared to their counterparts without tumor deposits in LNs (p=0.026, HR 123.93) and with lower proliferative indices (p=0.029, HR 79.68). These results reinforce the established role of LN metastasis and cellular proliferation rates as reliable indicators of relapse and survival in breast cancer patients [48–50], while also suggesting underlying molecular mechanisms that contribute to cancer recurrence [50]. This further underscores the importance of these factors in staging, prognosis, and therapy of invasive breast cancer in Sri Lankan patients.

One of the most paramount points in our study is the validation of the observed association between LN metastasis and RFS (p=0.026) by the compounded association of *PIK3CA* mutations with both LN metastasis (p=0.036) and significantly reduced RFS (p<0.001). This enables us to bridge all three findings. Additionally, the multivariate model demonstrated the compounded impact of all these factors (p<0.001) on the likelihood of disease relapse, ultimately emphasizing the importance of considering all three factors-*PIK3CA* mutations, LN status, and Ki67 index-along with patient ancestry, in prognosis and treatment planning for breast cancer patients in Sri Lanka and across South Asia. This may also suggest an underlying mechanistic link that contributes to the aggressive behavior of breast cancer. It is plausible that in ancestries more prone to the occurrence of *PIK3CA* mutations in malignant cells, these mutations promote the cancer spread via lymphatic channels, leading to LN metastasis and subsequent recurrence. This progression highlights a crucial step in cancer spread, as LNs function as critical filtering sites within the lymphatic system, where cancer cells infiltrate and establish secondary tumor deposits [52].

The above findings align with Stephen Paget’s ‘Seed and soil hypothesis,’ which proposes that metastasis occurs when certain tumor cells (the “seed”) preferentially grow in particular organ microenvironments (the “soil”) - and that metastases only develop when the right seed is implanted in the right soil [53]. Our study suggests that *PIK3CA* driver mutations, possibly the predominant H1047R mutation, may act as a genetic change that enhances the metastatic potential of breast cancer cells by promoting survival and proliferation. These genetic alterations could facilitate the invasion of lymphatic vessels, leading to the establishment of metastatic colonies in LNs. In this context, *PIK3CA* mutations may assist in planting the “seed” in the “soil” by promoting the establishment of tumor deposits in axillary LNs. Strikingly, further developments to this hypothesis, elaborate how the above mechanisms could result in the primary breast cancer metastasizing to the lungs and bones [54], the exact two organs of relapse diagnosed in our *PIK3CA*-mutated patient population. Therefore, by elucidating this molecular pathway, our study offers valuable insights into the potential role of *PIK3CA* in the underlying biology of aggressive breast cancers, particularly in the Sri Lankan population.

To our knowledge, PI3K inhibitors are not currently prescribed for breast cancer patients in Sri Lanka. Our study’s preliminary observation of a negative prognostic outcome in the predominantly SHR+/HER2-, Sri Lankan breast cancer cohort with *PIK3CA* mutations, serves as a calling out for Sri Lankan clinicians to incorporate *PIK3CA* mutational testing in routine diagnostic protocols. Patients identified with these mutations could benefit from PI3Kα inhibitors such as Alpelisib, which are approved for this clinical scenario and are known to prolong the RFS in patients [55]. Furthermore, this treatment approach could be tailored for high risk ethnic groups within Sri Lanka (i.e. those with Dravidian ancestry), should a larger study with a better representation of ethnic minority groups, confirm the association of *PIK3CA* mutations with patient ancestry. This personalized approach could extend beyond breast cancer to other malignancies such as colorectal, gastric, ovarian, and cervical cancers, which have also exhibited *PIK3CA* mutations and consequent impacts on prognosis [8,56].

Despite the significant implications of our study, there are several limitations. The primary limitation is the sample size, which was restricted due to financial and logistical constraints. This limitation impacted the statistical power of several results and affected the ability to perform robust regression and multivariate analyses. Additionally, the retrospective nature of our study also introduced certain constrains. For example, the grouping of patients into broad ancestral categories was based on self-reported ethnicity in hospital records of the patients, rather than genetic testing. Although studies indicate a high concordance between self-reported ancestry and genetic testing results [22], potential misclassification may have oversimplified the complex genetic landscape of the Sri Lankan population. Furthermore, the underrepresentation of minority ethnic groups due to the uneven geographical distribution of these populations, greatly impaired the power of the detected association of ancestry with mutation status. Overcoming this limitation might require recruiting patients from a broader array of hospitals and oncology clinics around the country, to ensure adequate representation of all ancestral groups.

Another limitation is the absence of data on other potentially interacting mutations within the *PIK3CA* gene or in other OGs and TSGs that were not tested in the cohort, and their interplay with the tested mutations. Additionally, the follow-up period in our study was relatively short. All patients with survival data (n=42) were alive, and 90.5% remained recurrence-free within 5 years post-diagnosis, which limited the survival analysis to RFS as a proxy. A longer follow-up period would provide a more comprehensive understanding of long-term outcomes. Nevertheless, this pilot study, the first to analyze *PIK3CA* mutations in relation to breast cancer in different ancestries in Sri Lanka, together with its observed prognostic implications, offers sufficient insight and direction to the chosen objectives, so that a larger, more conclusive study can be conducted to confirm and validate the reported findings. Accordingly, a study on a larger Sri Lankan cohort with a subsequent functional analysis is necessary to address the above limitations, especially to confirm the absence of the E545K mutation in Sri Lankan breast cancer patients and to validate the significance of observed mutations, particularly in the Dravidian population, which appears to be at higher risk based on our findings.

## Conclusion

This study is the first to report on *PIK3CA* mutations in Sri Lankan breast cancer patients, a population with a unique genetic heritage, demonstrating a *PIK3CA* mutation prevalence of 17.46% and significant associations between ancestry-linked mutations, LN metastasis and cancer relapse. Our findings also suggest the existence of a possible molecular link contributing to metastasis.

Notably, our observation of a significant association between patient ancestry and *PIK3CA* mutation status is the first of its kind in an exclusively South Asian cohort. This invites comparisons with similar findings - such as the absence of the E545K mutation - in global populations with shared ancestries. Our findings also provide a foundation for prognostic stratification and therapeutic decision-making for breast cancer patients in Sri Lanka - rooted in the presence of *PIK3CA* mutations and patient ancestry - for identifying high-risk patient subsets who may benefit from targeted therapies such as PI3K inhibitors, which are not currently prescribed in Sri Lanka. Further large-scale functional studies are necessary to validate these findings and to improve clinical outcomes for breast cancer patients in Sri Lanka.

## Data Availability

The data generated or analyzed in this study are included within the article and its supplementary data files. Raw data were generated and processed by the authors and are available upon reasonable request from the corresponding authors.

## Acknowledgements

We gratefully acknowledge the invaluable support provided by the staff at the Centre for Immunology and Molecular Biology (CIMB) and the Centre for Transgenic Technologies at the Faculty of Science, University of Colombo, Sri Lanka, as well as the medical laboratory technicians at the Diagnostics Laboratory of University Hospital KDU, Sri Lanka in the completion of the laboratory component of this research.

## Supporting Information

**S1 Fig. Ethical approval for the study granted by the Ethics Review Committee of the Institute of Biology, Sri Lanka (ERC IOBSL 303 07 2023)**

**S2 Fig. Age distribution of the cohort**

**S3 Fig. Distribution of tumor size (cm) in the cohort**

**S1 Table. Grouping of the variables used for the data analysis**

**S2 Table. Status of lymph node (LN) metastasis and patient age group in selected clinical and histopathological groups**

